# Assessment of physiological signs associated with COVID-19 measured using wearable devices

**DOI:** 10.1101/2020.08.14.20175265

**Authors:** Aravind Natarajan, Hao-Wei Su, Conor Heneghan

## Abstract

Respiration rate, heart rate, and heart rate variability are some health metrics that are easily measured by consumer devices and which can potentially provide early signs of illness. Furthermore, mobile applications which accompany wearable devices can be used to collect relevant self-reported symptoms and demographic data. This makes consumer devices a valuable tool in the fight against the COVID-19 pandemic. We considered two approaches to assessing COVID-19 – a symptom-based approach, and a physiological signs based technique. Firstly, we trained a Logistic Regression classifier to predict the need for hospitalization of COVID-19 patients given the symptoms experienced, age, sex, and BMI. Secondly, we trained a neural network classifier to predict whether a person is sick on any specific day given respiration rate, heart rate, and heart rate variability data for that day and and for the four preceding days. Data on 1,181 subjects diagnosed with COVID-19 (active infection, PCR test) were collected from May 21 – July 14, 2020. 11.0% of COVID-19 subjects were asymptomatic, 47.2% of subjects recovered at home by themselves, 33.2% recovered at home with the help of someone else, 8.16% of subjects required hospitalization without ventilation support, and 0.448% required ventilation. Fever was present in 54.8% of subjects. Based on self-reported symptoms alone, we obtained an AUC of 0.77 *±* 0.05 for the prediction of the need for hospitalization. Based on physiological signs, we obtained an AUC of 0.77 *±* 0.03 for the prediction of illness on a specific day with 4 previous days of history. Respiration rate and heart rate are typically elevated by illness, while heart rate variability is decreased. Measuring these metrics can help in early diagnosis, and in monitoring the progress of the disease.

## I. INTRODUCTION

The year 2020 has seen the emergence of a global pandemic caused by the Severe Acute Respiratory Syndrome Coronavirus 2 (SARS-CoV-2) virus. The disease caused by this virus typically presents as a lower respiratory infection, though many atypical presentations have been reported. This has caused a major health challenge globally due to the apparent high transmissibility of this virus in a previously unexposed population. Of particular concern is that the primary mechanisms by which the disease is transmitted are still somewhat under debate (e.g., the importance of airborne transmission)^1^, and the potential for infection by asymptomatic and pre-symptomatic patients (see for e.g., the discussion in Oran & Topol^2^). The disease is highly contagious, with transmission possible 2.3 days prior to the onset of symptoms, and peaking 0.7 days prior to the onset of symptoms according to one model^3^. As a result, a great deal of effort is underway to potentially diagnose COVID-19 early.

The popularity and widespread availability of consumer wearable devices has made possible the use of health metrics such as respiration rate, heart rate, heart rate variability, sleep, steps, etc in order to predict the onset of COVID-19 or similar illnesses. A 1^*◦*^C rise in body temperature can increase heart rate by 8.5 beats per minute on average^4^. Measuring the resting heart rate, or heart rate during sleep can therefore be a useful diagnostic tool. Similarly, the respiration rate is elevated when patients present with a fever^5^. Heart rate variability is the variability in the time between successive heart beats (the time between successive heart beats is called the “RR interval”), and is a valuable, non-invasive probe of the autonomic nervous system^6–8^. Lowered values are indicative of increased mortality^9^, and may provide early diagnosis of infection^10^. A study of heart rate variability in critically ill COVID-19 patients showed that the approximate entropy and the sample entropy were decreased in COVID-19 patients compared to critically ill sepsis patients^11^.

Zhu et al.^12^ studied heart rate, activity and sleep data collected from Huami wearable devices to potentially identify outbreaks of COVID-19, and concluded that at a population level an anomaly detection algorithm provided correlation with the measured infection rate. Menni et al.^13^ analyzed symptoms reported through a smartphone app and developed a model to predict the likelihood of COVID-19 based on the symptoms. Marinsek et al.^14^ studied data from Fitbit devices as a means for early detection and management of COVID-19. Miller et al.^15^ used the respiration rate obtained from Whoop devices to detect COVID-19. Mishra et al.^16^ analyzed heart rate, steps, and sleep data collected from Fitbit devices to identify the onset of COVID-19.

In this paper, we consider the correlation between changes in physiological signs related to respiration rate, heart rate and heart rate variability, and the corresponding presence of diseases assessed both through confirmed laboratory testing and self-reported symptoms and the time-course of the disease. We show that it is possible to use changes in these physiological metrics to detect illness, and provide estimates of sensitivity and specificity.

In addition, given the reporting of symptoms by study participants, we provide an estimate of predicted disease severity based solely on symptoms.

## II. MATERIALS AND METHODS

### A. Data Collection

Fitbit is a global leader in wearable technology since 2007, and has a large established base of users (over 30 million as of 2020). A significant percentage of its devices are configured to measure heart rate, and the underlying interbeat intervals (RR) that characterize heart rate variability. The Fitbit app provides a convenient user-facing app that can be configured to present user-facing questions, and to reliably capture responses in a secure and scalable way. In this study, active Fitbit users in the USA and Canada were invited to participate in a survey of whether they have experienced COVID-19 or similar infections, whether they had been tested, and to report on symptoms they experienced. They could also optionally provide additional demographic data such as age, sex, body mass index, and relevant background medical information such as underlying conditions such as diabetes, coronary arterial disease, or hypertension. While the researcher hypothesis was that metrics that could be generated from heart rate were likely to be most predictive of infectious disease, we did not restrict the survey to only Fitbit users with heart rate enabled devices (in practice 95% of survey respondents had heart rate enabled devices).

The survey and associated marketing and recruitment materials were approved by an Institutional Review Board (Advarra) and from May 21st 2020, the survey was available for completion by Fitbit users in the USA and Canada. The data presented here represent the analysis on survey results collected up to July 14, 2020. The survey contained the following questions in relation to COVID-19 and other likely confounding infectious diseases such as influenza, urinary tract infections etc. – a) have you been tested for COVID-19 (with separate sections provided for tests for active infection versus serological tests for previous infection), b) what were the symptoms experienced and the dates of onset and disappearance of the symptoms, c) were you tested for other infectious diseases such as influenza, strep throat etc.

Table I shows the overall breakdown of the survey responses as well as providing summary demographic information on the survey respondents. The age distribution of survey respondents was very similar to Fitbit users’ overall age distribution. Survey respondents skewed slightly more female than the corresponding figure for the overall general Fitbit population.

**TABLE I:** Breakdown of overall survey results by summary demographics and test responses

| *n*=132,454 completed survey |  |  |
| --- | --- | --- |
| **Overall Demographics** |  |  |
| Age (mean ± std) | 43.1 ± 13.9 | |
| Sex (M/F/Other) | 35,305/95,738/1,271 |  |
| **Active COVID Infection Tests (PCR)** |  |  |
|  | *n* | % of participants |
| Tests Reported | 16,231 | 12.25% |
| Positive | 1,181 | 0.89% |
| Negative | 13,662 | 10.31% |
| Awaiting Results/Unknown | 1,388 | 1.05% |
| **Serology Tests** |  |  |
|  | *n* | % of participants |
| Tests Reported | 8,044 | 5.12% |
| Positive | 625 | 0.47% |
| Negative | 6,780 | 5.12% |
| Awaiting Results/Unknown | 639 | 0.48% |
| **Flu Tests** |  |  |
|  | *n* | % of participants |
| Tests Reported | 3,085 | 2.33% |
| Positive Tests | 665 | 0.50% |
| Negative | 2,392 | 1.81% |
| Awaiting Results/Unknown | 27 | 0.02% |
| **Other Tested Infections** |  |  |
|  | *n* | % of participants |
| Tests Reported | 1,135 | 0.86% |

Table II describes the self-reported major comorbidities of the participants who reported a positive diagnosis of COVID-19, either through a confirmed PCR test or a serological test (we assume that the vast major-ity of tests for active COVID-19 infection were done using PCR-based techniques). Note that participants could also decline to answer this question, so these numbers are only indicative of general trends in the disease population.

**TABLE II:** Self-reported health characteristics of participants who reported either a positive PCR test or a positive serological test. Note that some participants may have received both.

| **Characteristics of COVID-19 Participants** |  |  |
| --- | --- | --- |
| Age (mean ± std) | 41.9 ± 12.9 yrs | |
| Sex (M/F/Other) | 410/1,223/9 |  |
| **Self Reported Health** |  |  |
|  | *n* | % |
| Hypertension | 294 | 17.9% |
| Asthma | 303 | 18.5% |
| Diabetes | 130 | 7.9% |
| Coronary Arterial Disease | 22 | 1.3% |
| Stroke | 19 | 1.2% |
| Chronic Lung Disease | 21 | 1.3% |
| Chronic Kidney Disease | 14 | 0.9% |
| Congestive Heart Failure | 12 | 0.7% |

**TABLE III:**
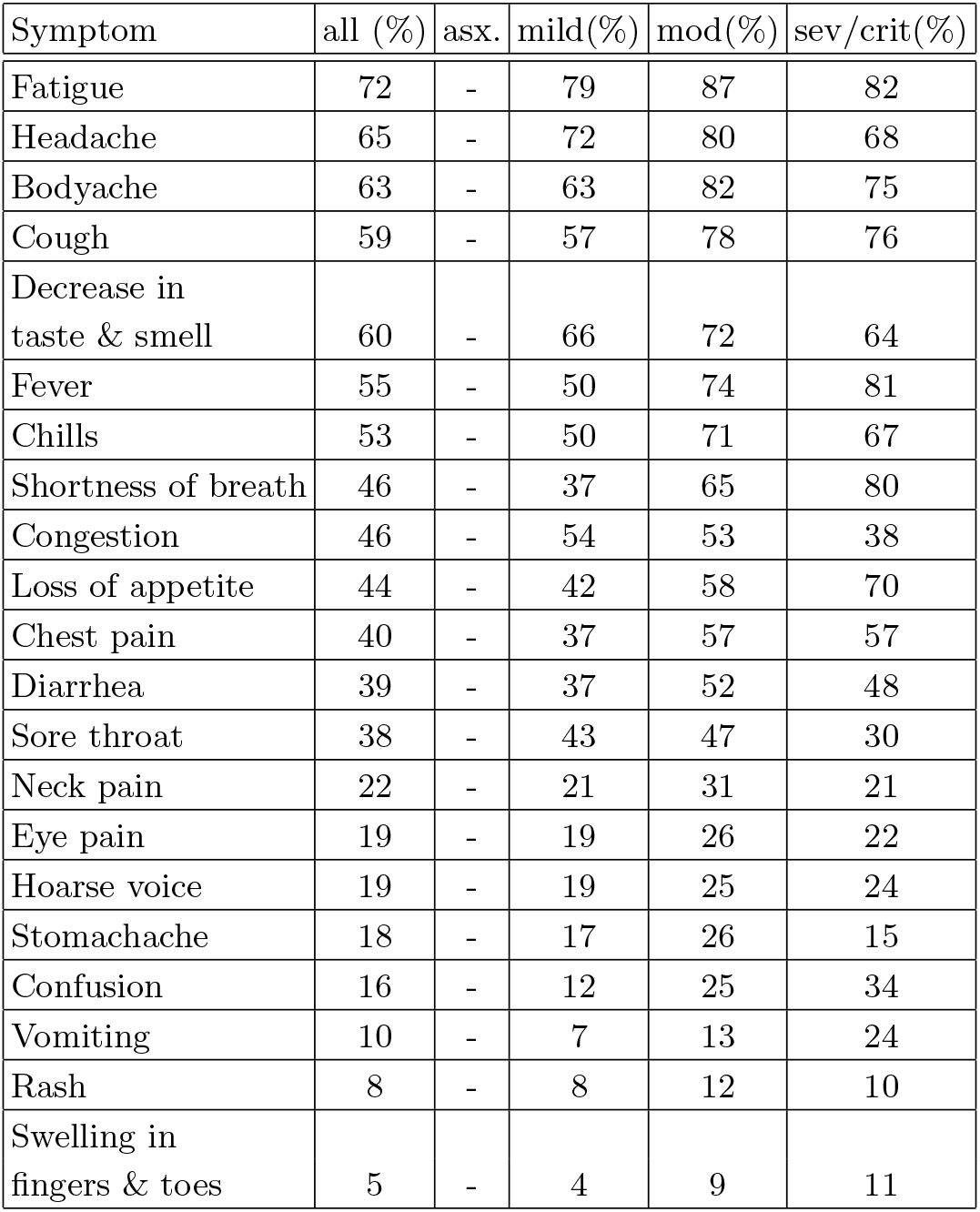
Prevalence of symptoms

In order to assess some metric of disease severity, we took the approach of asking about the person’s treatment rather than through quantification of symptoms. The options provided to a survey participant were:

1. I didn’t experience symptoms.
2. I self-treated alone.
3. I self-treated with someone’s help.
4. I required hospitalization without ventilation support.
5. I required ventilation.
6. Prefer not to say.

We consider 1 as asymptomatic. 2 is assigned to category “mild”. 3 is assigned to “moderate”, 4 is “severe” and 5 is “critical”. 11.0% of participants were asymptomatic, 47.2% had mild symptoms, 33.2% were moderate, 8.16% severe, and 0.448% critical. Those with mild symptoms recovered sooner than those with moderate or severe symptoms. The distribution in the duration of symptoms is shown in Fig. 1.

**FIG 1:**
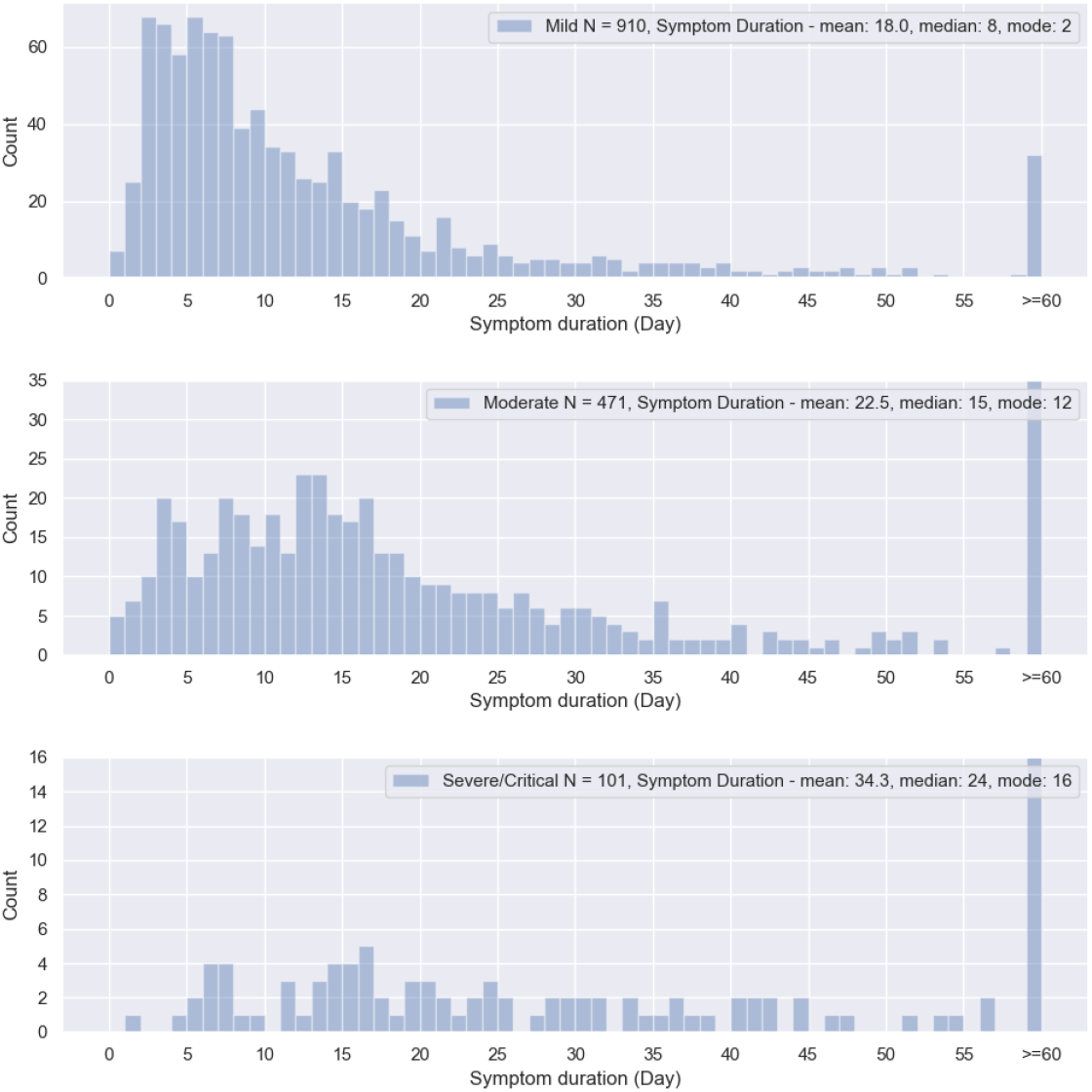
Distribution of symptom duration for mild, moderate and severe/critical cases. The median symptom duration is 8 days for mild cases, 15 days for moderate cases, and 24 days for severe/critical cases.

### B. Making predictions based on health metrics and symptoms

Firstly, we wished to use the self-reported symptoms as a classifier of likely need for hospitalization. Using the symptoms along with the age, sex, and BMI as input features, we trained a logistic regression classifier to predict the need for hospitalization. We only considered symptomatic individuals for this analysis.

Secondly, we considered the use of physiological signs to predict the presence of symptoms associated with COVID-19. Let us denote the *n*^th^ day relative to the start of symptoms as *D_n_. D*_0_ thus represents the day when symptoms started. We make the assumption that individuals are healthy, i.e. class “Negative” from day *D_a_* to day *D_b_* where *D_b_ < D*_0_. The days between *D_b_* and *D*_0_ are treated as a buffer space when subjects may or may not be sick, and hence, ignored. Subjects are considered to be sick from day *D_c_* up to day *D_d_* where *D_c_ ≥ D*_0_. The choices of *D_a_, D_b_, D_c_*, and *D_d_* are made through cross validation. As a guide to choosing the days, we note the median incubation period is estimated at 5.1 days^17^. Since we are using the date relative to the start of symptoms as the ground truth label, we consider only symptomatic individuals.

The following physiological data was calculated for each user on a daily basis using the data recorded from their Fitbit device:

- The estimated mean respiration rate during deep (slow wave) sleep (we default to light sleep if deep sleep data is insufficient).
- The mean nocturnal heart rate during non-Rapid Eye Movement (NREM) sleep.
- The Root Mean Square of Successive Differences (RMSSD) of the nocturnal RR series.
- The Shannon entropy of the nocturnal RR series.

Data are collected simultaneously from the PPG sensor and the accelerometer. RR data are only stored when no motion above a set threshold is detected, and when the coverage in a 5 minute window exceeds 70%. Data are only collected when the subjects are at rest. The RR data are then cleaned to remove noise due to missed heart beats, motion artifacts, electronic noise, etc. The Fitbit system estimates periods of light, deep (slow wave) and REM sleep^18^ and this is used in deciding which sections of the overnight data to process. The respiration rate is obtained by fitting a Gaussian model to the spectrum of the interpolated RR intervals as a function of frequency - this relies on the phenomenon of respiratory sinus arrhythmia (RSA) to induce a measurable modulation of the RR interval series. In cases where there is no discernible RSA, we do not estimate the respiration rate. The RMSSD is a time domain measurement used to estimate vagally mediated changes^8^. It is computed in 5 minute intervals, and the median value of these individual measurements over the whole night is calculated. The Shannon entropy is a non-linear time domain measurement computed using the histogram of RR intervals over the entire night. The RMSSD and entropy are computed between midnight and 7 am. The sleeping heart rate is estimated from non-REM sleep only. The respiration rate is computed from deep sleep when possible, and from light sleep in the case of insufficient deep sleep.

Since health metrics such as respiration rate, heart rate, and heart rate variability can vary substantially between users, we use the *Z-*scored equivalents:

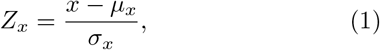

where *x* could stand for respiration rate, heart rate, RMSSD, or entropy. µ_x_ and σ_x_ are the rolling mean and rolling standard deviation of the metric being measured. For each day *D_n_*, we construct a 5 × 4 matrix with the normalized Z-scores corresponding to the 4 health metrics, measured on days *D_n_ & D_n_*-4. Each day is represented by a matrix with that day's data along with the previous four days data. Each row of the matrix represents a day of data, while each column represents a metric. We linearly interpolate missing data, but only do so if there is a minimum of 3 days of data. We create an “image” from each matrix by resizing each 5 × 4 matrix to a 28×28×1 matrix, with the last dimension indicating that there is only one color channel. The pixel values are rescaled to the range (0,1). We included 673 symptomatic individuals with sufficient data for analysis. 70% of the subjects were randomly selected to comprise the training set. The remaining 30% of subjects were split equally into 2 hold-out sets: one for cross-validation, and one for testing.

Fig. 2 shows the neural network architecture. Each image is input to a 1-dim. convolutional stage with *m* filters, and a filter size of *k*. After maxpooling, the convolutional stage produces a set of *m* features. Non-linearity in the form of a “Relu” layer is introduced. A dense layer is used to reduce the *m* convolutional features to a small feature set *N*_1_. At this stage, an array of *n* external inputs is applied including features such as age, gender, and BMI that need to bypass the convolutional stage. We also input the sleep efficiency for day *D_n_* defined as the ratio of time asleep to the duration between wake time and bed time. In addition, we also include the *Z-*scored respiration rate, heart rate, RMSSD, and entropy for day *D_n_* as a part of the array. The final dense layer leads to a softmax layer with two possible output classes.

**FIG 2:**
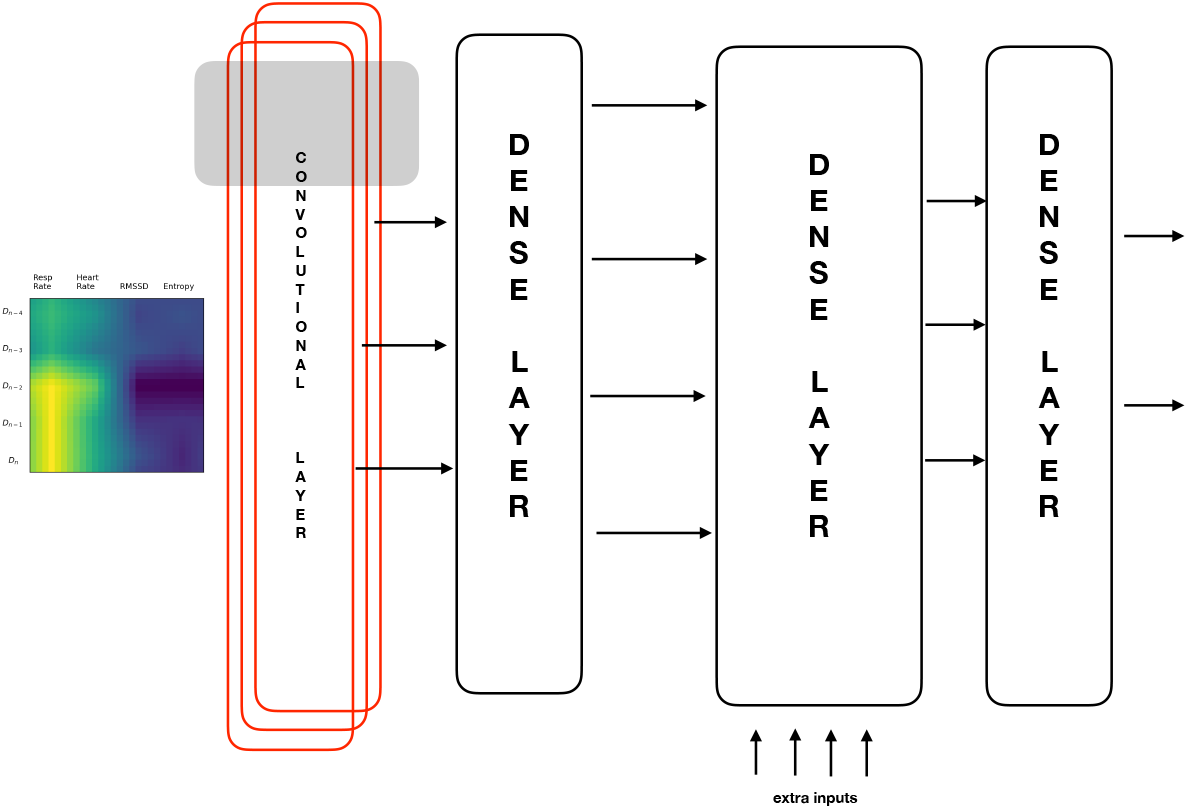
The neural network architecture. The nocturnal respiration rate, heart rate, RMSSD, and entropy for day *D_n_* along with the previous four days data are *Z*-scaled, arranged in the form of a 5*×*4 matrix and rescaled to 28 × 28 × 1. This image is fed to a 1-dim. convolutional layer with *m* filters. The first dense layer reduces these *m* features to a smaller number of *N*_1_ features which are concatenated with an array of external inputs such as age, gender, BMI, etc. The last dense layer leads to a softmax filter.

## III. RESULTS

Firstly we report on the prediction of hospitalization based on symptoms. We trained a logistic regression model to predict the need for hospitalization with four fold cross-validation, using the symptoms as input features, along with the age, sex, and BMI. Fig. 3 shows the ROC curve where “true positive” indicates a prediction of hospitalization for an individual who indeed required hospitalization. Averaged over four folds, the Area Under ROC (AUC) is 0.77 *±* 0.05. The probability of the need for hospitalization *p* may be expressed as:

**FIG 3:**
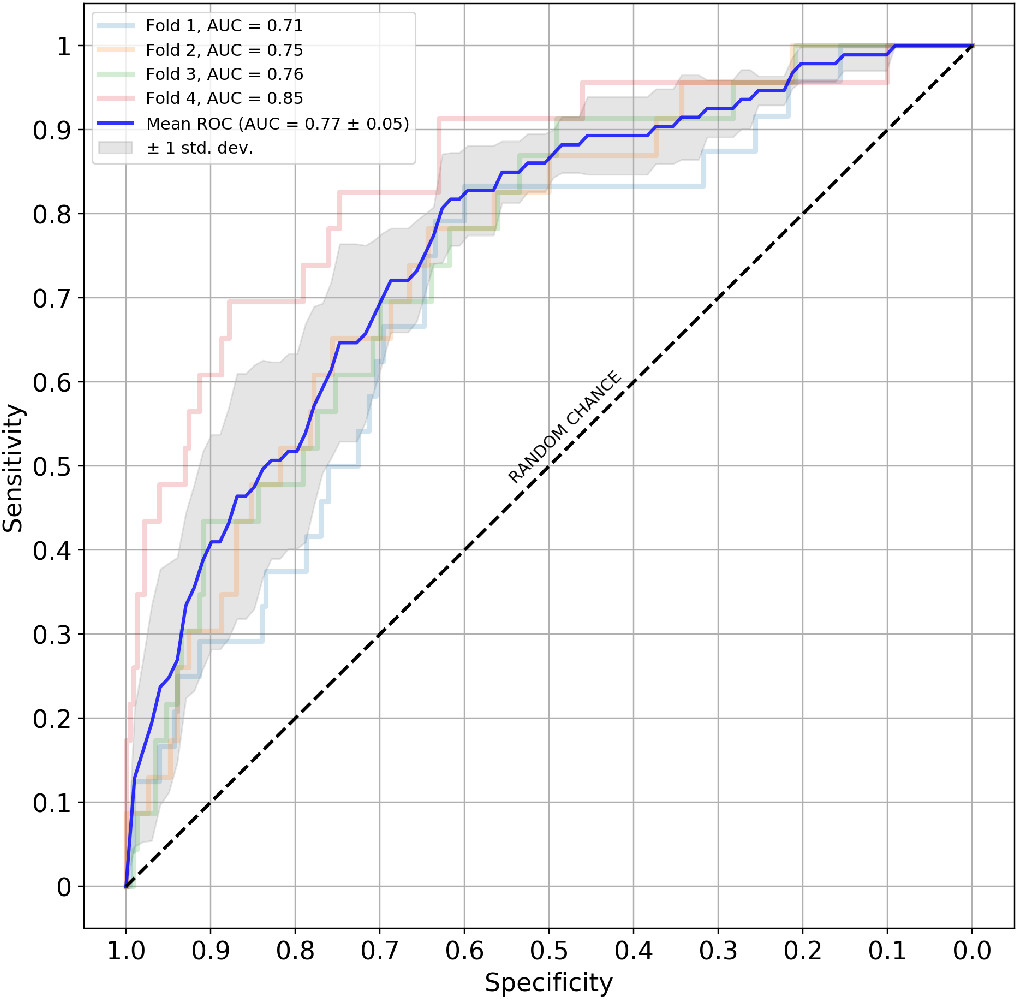
Predicting the need for hospitalization given the symptoms. The AUC averaged over 4 folds is 0.77 *±* 0.05.

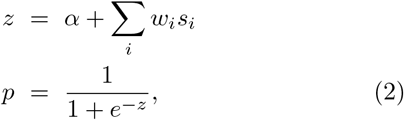

where *α* = *−*3.3457, *s_i_* is a symptom (1 if the symptom is present, and 0 otherwise), and *w_i_* is the weight corresponding to symptom *s_i_*. The weights for the various symptoms are shown in Table IV. We rescale the 2 continuous variables (age and BMI) as follows:

**TABLE IV:** Predicting the need for hospitalization: Importance of symptoms.

| Symptom | Weight |
| --- | --- |
| Shortness of breath | 1.1210 |
| Vomiting | 1.0784 |
| Loss of appetite | 0.7156 |
| Confusion | 0.6442 |
| Swelling in the fingers and toes | 0.4670 |
| Hoarse voice | 0.4520 |
| Fever | 0.4374 |
| age (scaled) | 0.3558 |
| BMI (scaled) | 0.3488 |
| Chest pain | 0.2730 |
| Cough | 0.1553 |
| Sex (male: yes/no) | 0.0750 |
| Chills | 0.0654 |
| Body ache | 0.0517 |
| Diarrhea | 0.0441 |
| Decrease in taste and smell | -0.1661 |
| Head ache | -0.2261 |
| Eye pain | -0.2282 |
| Congestion | -0.3258 |
| Neck pain | -0.3739 |
| Rash | -0.4046 |
| Fatigue | -0.4651 |
| Sore throat | -0.7454 |
| Stomach ache | -0.7575 |

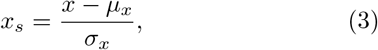

where *x_s_* is the scaled version and *x* stands for age or BMI. *µ_x_* and *σ_x_* are the respective mean and standard deviation, which are (41.5 years, 12.5 years) for age and (30.8, 7.7) for BMI. For the sex, 1 indicates male and 0 indicates female.

Let us now consider the problem of determining whether an individual is sick or healthy given the physiological metrics. Fig. 4 show the average *Z*-scores of symptomatic individuals for the respiration rate, heart rate, RMSSD, and entropy as a function of day, where day *D*_0_ represents the start of symptoms. The error bars represent the standard error of the mean. The respiration rate shows the largest effect and also takes the longest time to return to its base value. The duration between day *≈ D*_+7_ and *≈ D*_+21_ may be thought of as a recovery phase during which the health metrics return to their normal values. It is interesting to note that the heart rate decreases on average, following day *D*_+7_, and returning to the base value by day *D*_+20_. The HRV metrics on the other hand, are slightly elevated on average in the recovery phase. We did not notice a decrease in respiration rate on average, during this phase.

**FIG 4:**
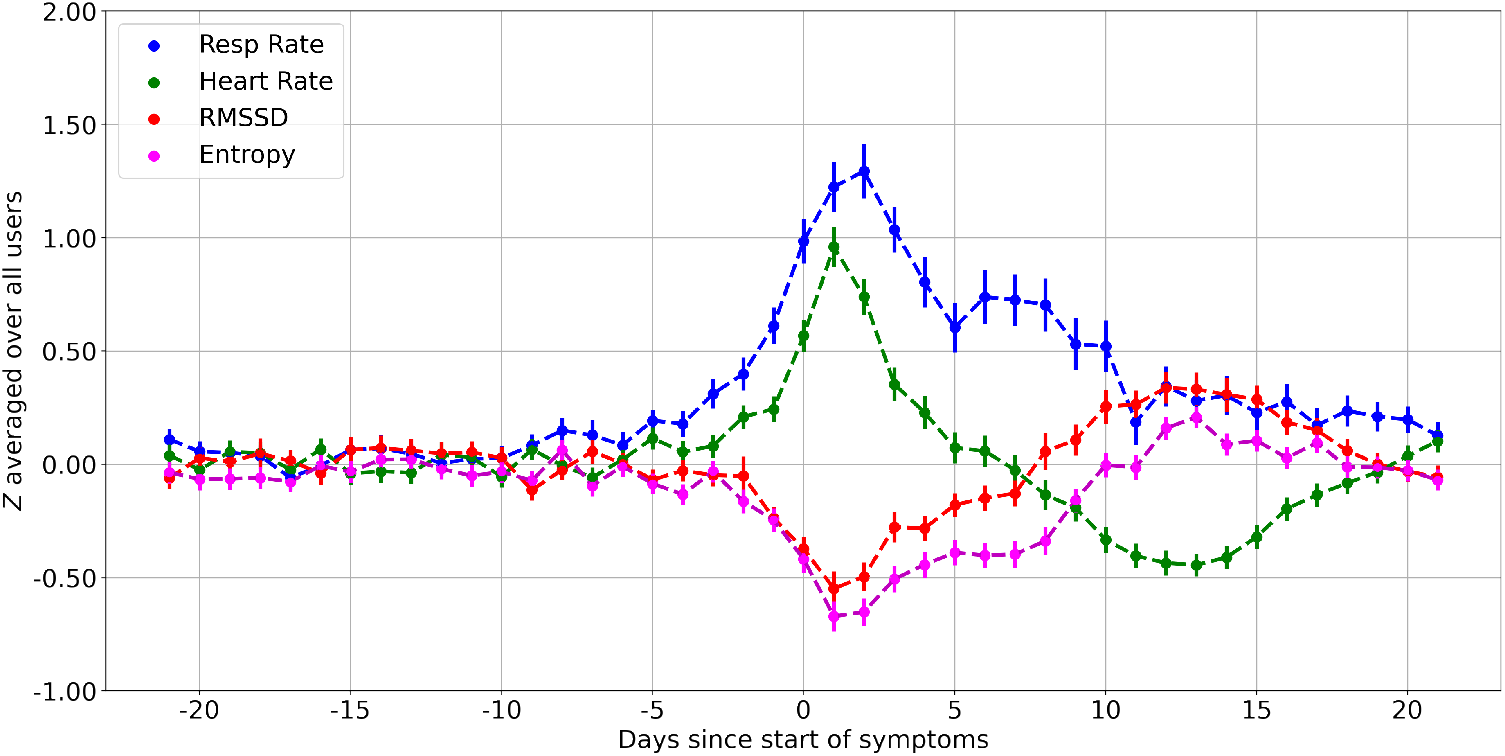
*Z*-scores for respiration rate, heart rate, RMSSD, and entropy. Day 0 (*D*_0_) represents the start of symptoms. The respiration rate and heart rate are elevated during times of sickness, while the RMSSD and entropy are decreased. These metrics may change a few days prior to the start of symptoms. The duration from day *D*_+7_ to day *D*_+21_ may be thought of as a recovery period during which the heart rate is slightly below the normal value, while the HRV is slightly elevated. The respiration rate takes the longest time to return to its base value, and does not show a recovery phase unlike the heart rate or HRV. Shown are the mean and standard error of the mean.

We trained a convolutional neural network to predict whether an individual is sick on any specific day given the *Z−*scores for respiration rate, heart rate, RMSSD, and entropy for that day and the preceding four days. Using the cross-validation set, we obtained the best performance considering negative classes from day *D_−_*_21_ to day *D_−_*_8_. The days from *D_−_*_7_ to *D*_0_ are discarded. Data for the positive classes come from day *D*_+1_ to day *D*_+7_. The filter size *k* for 1-dimensional convolution was set to 5 pixels, while the number of filters *m* was set to 64. A drop-out of *p* = 0.4 was applied for regularization. The number of neurons in the first dense layer *N*_1_ = 12, while the number of neurons in the second dense layer *N*_2_ = 64. The data are randomly split into training and hold-out sets, but we do this four times using a different random seed each time, to reduce the risk of outliers influencing the results. The AUC for the four folds was found to be 0.81, 0.74, 0.78, and 0.74. The mean AUC over 4 folds is 0.77 *±* 0.027. At 95% specificity, the sensitivity averaged over 4 folds is 44% *±* 2.3%.

Having set the network parameters, we retrained the classifier using all available data up to June 1, 2020. Data after June 1 was set aside a hold-out set for testing. On this dataset, the AUC was found to be 0.80, with a sensitivity of 47% at a specificity of 95%. Fig. 6 shows the fraction of symptomatic users who are classified as sick, for different days. Day 0 is the start of symptoms. Negative numbers indicate days prior to the start of symptoms, while positive numbers are days following the start of symptoms. Plot (a) shows the results for 95% specificity (solid, blue curve) and 90% specificity (dashed, green curve). Plot (b) shows the results for subjects at 95% specificity, who present with specific symptoms such as a fever, cough, or congestion. Plot (c) shows the effect of disease severity on the probability of illness detection, with a specificity of 95%.

**FIG 5:**
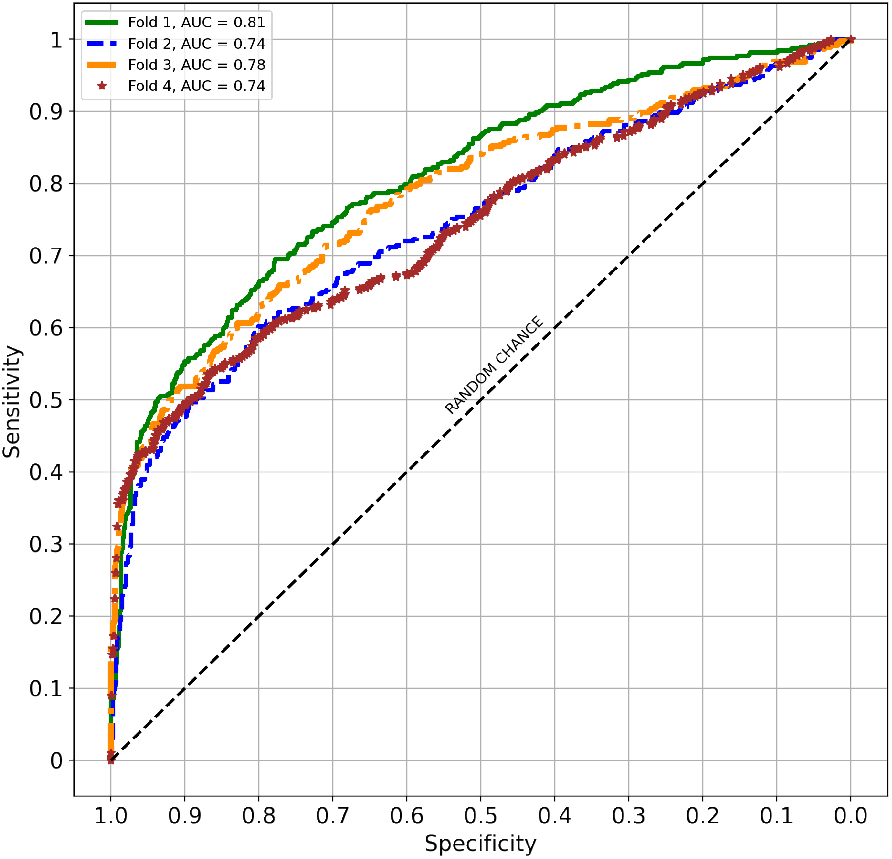
Predicting sickness given the physiological signs. With 4 fold validation, the AUC is 0.77 *±* 0.027. At 95% specificity, the sensitivity averaged over 4 folds is 44% *±* 2.3%.

**FIG 6:**
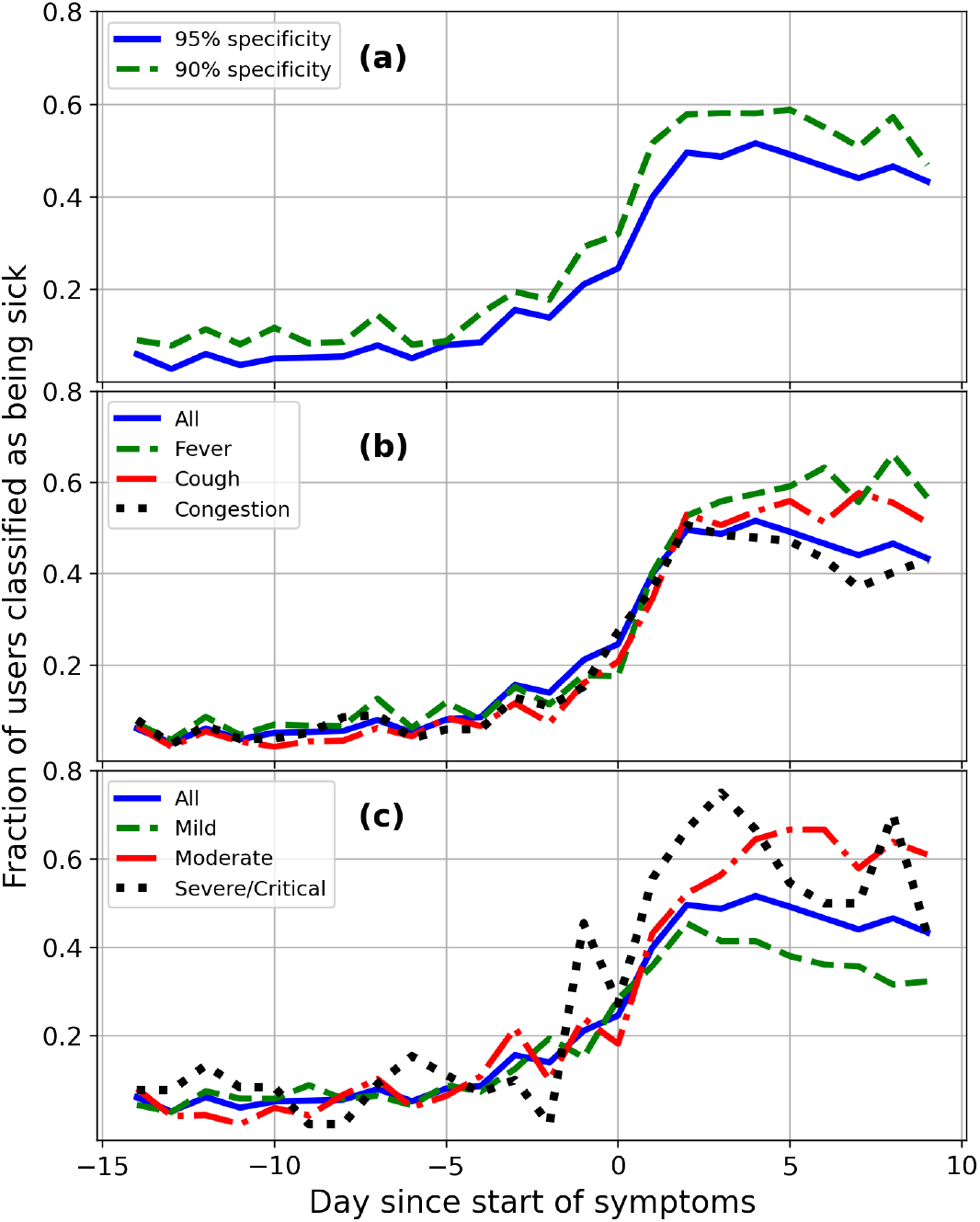
Fraction of users in the test set who predict positive. The solid, blue curve in (a) shows the predictions for a 95% specificity scenario, while the dashed, green curve shows the same, for 90% specificity. Plot (b) shows the same predictions (95% specificity) but for subjects who present with specific symptoms. The presence of a fever (dashed, green) or a cough (dot-dashed, red) increases the probability of detection more than some other symptoms such as congestion (dotted, black). Plot (c) shows the same curves (95% specificity), but for different severity. As expected, severe (dotted, black) and moderate (dot-dashed, red) cases are easier to detect than mild (dashed, green) cases.

## IV. DISCUSSION

In this article, we analyzed data on 1,181 subjects diagnosed with COVID-19 using the active infection PCR swab test, in the time period May 21 – July 14, 2020. All subjects wore Fitbit devices and resided in the United States or Canada. The overall positivity rate was 7.26%. 11.0% of reported cases were asymptomatic. 8.16% of subjects required hospitalization without ventilation support, while 0.448% required ventilation. Fatigue was the most common symptom, present in 72% of cases. Symptoms such as rash and swelling in the fingers and toes presented rarely in less than 10% of cases.

The duration of symptoms depends on the severity: Mild cases show a median duration of 8 days, while moderate cases have a median duration of 15 days. The median duration for cases that required hospitalization was found to be 24 days with a large spread, with several cases with duration exceeding 2 months. We provided a simple formula to estimate the need for hospitalization given the symptoms, age, sex, and BMI. Shortness of breath is highly indicative of the need for hospitalization, while sore throat and stomach ache were the least likely. Rather surprisingly, gastrointestinal symptoms such as vomiting and loss of appetite were indicative of severe illness. Among demographic information, being older and having a high BMI show higher likelihoods for hospital-ization.

We showed that respiration rate, heart rate, and heart rate variability are useful indicators of the onset of illness. We trained a convolutional neural network to predict illness on any specific day given health metrics for that day and the preceding four days. With 95% specificity, this classifier can detect 40% of cases on day *D*_+1_, where *D*_0_ is the date when symptoms present. The classifier can detect 25% of cases on day *D*_0_, and 21% of cases on day *D_−_*_1_. For the 90% specificity case, the corresponding numbers are 52% on day *D*_+1_, 32% on day *D*_0_, and 29% on day *D_−_*_1_. The peak values for the 95% (90%) specificity scenarios are 52% (59%) on day *D*_+4_ (day *D*_+5_). Considering only subjects who present with a fever, we classify 63% of subjects as positive, on day *D*_+6_, with 95% specificity. The fraction who are classified positive increases with increased severity, but this effect is pronounced several days after the start of symptoms. Considering only subjects who show a moderate/severe/critical presentation, 65% of subjects are classified positive on day *D*_+4_, with 95% specificity.

This study has multiple limitations which may confound some of its findings. The survey participants were all Fitbit users which may not represent the general US and Canadian population, and were all self-selecting in responding to the survey. Participants were asked to self-recall the start-date and end-date of any symptoms they experienced which may be quite unreliable. Participants may also confuse active (PCR) tests with serological (antibody) tests. In order to simplify the survey, we did not ask for a breakdown of symptom presentation and severity through out the time-course of the disease. For the prediction of the need for hospitalization, our data consisted of 95 positive cases and 926 negative cases, so it is possible that our results are potentially affected by the small number of positive cases, as well as by the class imbalance.

Nevertheless, we believe this survey provides an important scientific contribution by suggesting (a) hospitalization risk can be calculated from self-reported symptoms, and (b) relevant and predictive physiological signs related to COVID-19 may be detected by consumer wearable devices.

## Data Availability

Data were collected under the guidance of an IRB and consistent with Fitbit's terms and conditions. We are unable to open source the data.

## Acknowledgments

We thank members of the Fitbit Research team and our collaborators at Scripps Translational Institute and Stanford University for illuminating discussions on COVID-19 and related topics. We thank the Fitbit users who volunteered their data for inclusion in this study.

